# Design and validation of HIV peptide pools for detection of HIV-specific CD4^+^ and CD8^+^ T cells

**DOI:** 10.1101/2022.04.29.22274468

**Authors:** Rita Alkolla, Alba Grifoni, Shane Crotty, Alessandro Sette, Sara Gianella, Jennifer Dan

**Affiliations:** Center for Infectious Disease and Vaccine Research, La Jolla Institute for Immunology, La Jolla, California, USA; Department of Medicine, Division of Infectious Diseases and Global Public Health, University of California, San Diego, La Jolla, California, USA; Consortium for HIV/AIDS Vaccine Development (CHAVD), The Scripps Research Institute, La Jolla, CA, USA

## Abstract

Reagents to monitor T cell responses to the entire HIV genome, based on well characterized epitopes, are missing. Evaluation of HIV-specific T cell responses is of importance to study natural infection, and therapeutic and vaccine interventions. Experimentally derived CD4^+^ and CD8^+^ HIV epitopes from the HIV molecular immunology database were developed into Class I and Class II HIV megapools (MPs). We assessed HIV responses in persons with HIV pre combined antiretroviral therapy (cART) (n=17) and post-cART (n=18) and compared these responses to 15 controls without HIV (matched by sex, age, and ethnicity). Using the Activation Induced Marker (AIM) assay, we quantified HIV-specific total CD4^+^, memory CD4^+^, circulating T follicular helper, total CD8^+^ and memory CD8^+^ T cells. We also compared the Class I and Class II HIV MPs to commercially available HIV gag peptide pools. Overall, HIV Class II MP detected HIV-specific CD4^+^ T cells in 21/35 (60%) HIV positive samples and 0/15 HIV negative samples. HIV Class I MP detected an HIV-specific CD8^+^ T cells in 17/35 (48.6%) HIV positive samples and 0/15 HIV negative samples. Our innovative HIV MPs are reflective of the entire HIV genome, and its performance is comparable to other commercially available peptide pools. Here, we detected HIV-specific CD4^+^ and CD8^+^ T cell responses in people on and off cART, but not in people without HIV.

## Introduction

The Human Immunodeficiency Virus (HIV) infects CD4^+^ T cells and, if unchecked, slowly obliterates the host immune response over time [1]. During initial infection, the host mounts an HIV-specific immune response with the development of HIV-specific CD4^+^ and CD8^+^ T cells [2–4]. With combined antiretroviral treatment (cART), HIV RNA levels in blood fall below the limit of detection, and the host immune system is “restored” with a rebound in CD4^+^ T cell count [5,6]. The quality of HIV-specific CD4^+^ and CD8^+^ T cells depends on if these individuals are elite controllers or chronic progressors.

To understand the HIV host immune response, it is necessary to precisely identify and quantify HIV-specific T cells. Classic methodologies to detect HIV-specific CD4^+^ and CD8^+^ T cell responses have relied on intracellular cytokine staining, Major Histocompatibility Complex (MHC) tetramer binding assays, lymphoproliferative assays, and ELISpots [7]. Intracellular cytokine staining requires fixation of cells which cannot be used in subsequent functional assays or most gene expression analyses. MHC tetramer binding assays require foreknowledge of an individual’s Human Leukocyte Antigen (HLA). Both lymphoproliferative assays and ELISpots require long term cultures during which cell phenotypes might change [8]. We therefore developed the Activation Induced Marker (AIM) assay to directly detect antigen-specific CD4^+^ T cells after a 24 hour cell culture [8–11]. The AIM assay is cytokine-independent and detects antigen-specific cells utilizing surface co-expression of specific activation markers, e.g. OX40, PD-L1, CD25, CD69, CD40L, and 41BB (CD137). Advantages of the AIM assay include (1) the ability to sort live cells by flow cytometry for subsequent assays or sequencing, (2) a short assay duration which limits detection of bystander activated cells, and (3) the ability to further classify antigen-specific T cells into different T cell memory subsets and T helper cell functions. For human lymphocytes, marker combinations of CD25/OX40, OX40/PD-L1, OX40/41BB, OX40/CD40L, CD69/CD40L, and CD69/41BB have been used to detect antigen-specific CD4^+^ and CD8^+^ T cells [8,10,12–14].

The HIV proteins gag and env are major targets of HIV-specific CD4^+^ and CD8^+^ T cell responses [2,16–18]. Prior studies have demonstrated a correlation of gag-specific CD4^+^ T cells and viremic control, whereas the reverse have been observed with env-specific CD4^+^ T cells [16]. However, gag and env do not reflect the entirety of the host immune response to HIV viral proteins. Herein, we generated a Class I HIV-specific megapool (MP) and Class II HIV MP, reflective of the entire HIV proteome. Using the HIV molecular immunology database to extract experimentally derived HIV epitopes and Immune Epitope Database (IEDB) cluster tool [19], we designed a Class I HIV MP and Class II HIV MP consisting of immunodominant peptides from env (envelope), gag, gp160 (polyprotein of env), nef, pol, rev, tat, vif, vpr, and vpu. We show that the Class I and Class II HIV MP are able to detect HIV-specific CD8^+^ and CD4^+^ T cells, respectively.

## Materials and Methods

### Study subjects

Peripheral blood mononuclear cells (PBMCs) were collected from people with HIV (PWH) pre-cART and post-cART (Table 1) between 2001 and 2018. These 21 study participants were enrolled under the San Diego Primary Infection Research Consortium (PIRC) (https://www.pirc.ucsd.edu) [20]. These individuals were diagnosed early in their HIV infection and were followed longitudinally before and after cART initiation. Participants were recruited under a protocol approved by the Institutional Review Board (IRB) at the University of California, San Diego (UCSD).

**Table 1.** Clinical Characteristics of HIV positive patients.

| **Pre-cART (n=17)** | **[Median, IQR]** |
| --- | --- |
| **Age (Years)** | 39 [29 – 43] |
| **Time from <sup>a</sup>EDI (days)** | 98 [74 – 2004] |
| **CD4 (cells/mm<sup>3</sup>)** | 606 [415 – 718] |
| **HIV viral load (copies/mL)** | 120,000 [55,087 – 244,480] |
| **Gender** |  |
| Male (%) | 100% |
| **Ethnicity** |  |
| Hispanic (%) | 11.8% |
| Non-Hispanic (%) | 88.2% |
| **Race** |  |
| White (%) | 82.3% |
| Black (%) | 5.9% |
| Other (%) | 11.8% |
| **Sexual Orientation** |  |
| MSM (%) | 100% |
| **Post-cART (n=18)** | **[Median, IQR]** |
| **Age (Years)** | 40 [33 – 46] |
| **Time from EDI (days)** | 983 [672 – 2838] |
| **Time from cART (days)** | 667 [495 – 1388] |
| **Time from cART to undetectable HIV viral load (days)** | 236 [162 – 644] |
| **CD4 (cells/mm<sup>3</sup>)** | 825 [563 – 1025] |
| **HIV viral load (copies/mL)** | 50 [45 – 50] |
| **Gender** |  |
| Male (%) | 100% |
| **Ethnicity** |  |
| Hispanic (%) | 22.2% |
| Non-Hispanic (%) | 77.8% |
| **Race** |  |
| White (%) | 77.7% |
| Black (%) | 5.6% |
| Other (%) | 16.7% |
| **Sexual Orientation** |  |
| MSM | 100% |

<sup>a</sup>EDI: estimated date of infection

Blood specimens were taken from 11 individuals at an early time point post infection (21-138 days post the estimated date of infection). Nine of the 11 individuals had paired pre- and post-cART specimens. One individual had only a pre-cART specimen and one individual had only a post-cART specimen.

Blood specimens were taken from 10 individuals who were chronically infected (441-3598 days post the estimated date of infection). Five of the 10 individuals had paired pre- and post-cART specimens. Two individuals had only a pre-cART specimen and three individuals had only a post-cART specimen.

As a control group, we used PBMCs from 15 HIV seronegative donors matched by sex, age, and ethnicity. Participants for the control group were recruited under a protocol approved by IRB at La Jolla Institute for Immunology (LJI).

### HIV Megapools (MP)

HLA Class I and II restricted epitopes have been extracted from HIV molecular immunology database (https://www.hiv.lanl.gov/content/immunology/) using the T Helper/CD4^+^ Search and the CTL/CD8^+^ search [21]. The list of epitopes spanning all the HIV proteins (env, gag, gp160, nef, pol, rev, tat, vif, vpr, vpu) have been then clustered using the Epitope Cluster Analysis Tool 2.0 (http://tools.iedb.org/main/analysis-tools/) via the cluster-break method with a 70% homology cutoff, as previously described for other megapool generation [19,22]. The corresponding list of HIV peptides were synthesized by TC Peptide Lab (San Diego) as crude material on 1-2 mg scale. Individual peptides were resuspended in dimethyl sulfoxide (DMSO), pooled, and sequentially lyophilized, as previously reported. The resulting lyocake was resuspended in DMSO at a stock concentration of 1mg/mL. The Class I MP consists of 187 peptides (46 gag peptides, 34 gp160 peptides, 25 nef peptides, 59 pol peptides, 4 rev peptides, 2 tat peptides, 9 vif peptides, 6 vpr peptides, and 2 vpu peptides). The Class II MP consists of 164 peptides (7 env peptides, 5 gag peptides, 61 gp160 peptides, 15 nef peptides, 61 pol peptides, 2 rev peptides, 4 tat peptides, 1 vif peptides, 7 vpr peptides, and 1 vpu peptide). Peptides are listed in Supplemental Tables 1 and 2.

### Other Antigens

CMV Class I and II peptide MPs were synthesized by TC Peptide Lab (San Diego) as crude material on 1-2 mg scale. Individual peptides were resuspended in DMSO and pooled into two different MPs for this study. The peptide pools were sequentially lyophilized and each pool resuspended at a stock concentration of 1mg/mL [8,23]. JPT HIV-1 gag peptide pool (JPT PepMix HIV (GAG) Ultra), consisting of 150 overlapping peptides of 15-mers with 11 amino acid overlap, was purchased from JPT Peptide Technologies. National Institutes of Health (NIH) HIV-1 Clade B gag peptide pool (NIH AIDS Reagent Program #12425), consisting of 123 overlapping peptides of 15-mers with 11 amino acid overlap, was provided by the AIDS Reagent Program. Each peptide pool (JPT and NIH) was suspended in DMSO at a final concentration of 1mg/mL.

### PBMC Isolation

PBMCs were isolated from whole blood by density gradient centrifugation using Histopaque 1077 (Sigma) and cryopreserved in liquid nitrogen in fetal bovine serum containing 10% DMSO.

### AIM Assays

Cells were cultured in RPMI supplemented with 5% human AB serum (Gemini Bioscience), Glutamax (Gibco), and Penicillin/Streptomycin (Gibco). Cryopreserved PBMCs were thawed and cultured at ∼ 1×10^6^ cells per well in a 96 well round bottom plate. Cells were stimulated with peptides: HIV Class 1 MP (1μg/mL), HIV Class II MP (2μg/mL), HIV JPT gag peptide pool (2μg/mL), HIV NIH gag peptide pool (2μg/mL), CMV Class 1 MP (1μg/mL), and CMV Class II MP (2μg/mL). As a positive control, cells were stimulated with SEB (100ng/mL, Toxin Technology). As a negative control, cells were stimulated with media containing an equivalent volume of DMSO to the one used for the peptide stimulation. When cells were limiting, priority stimulations were HIV Class II MP > HIV Class I MP and CMV Class II MP > CMV Class I MP. Cells were cultured for 24 hours and stained by flow cytometry (FACS). FACS staining buffer consisted of 0.5% Bovine serum albumin (BSA) in phosphate buffered saline (PBS).

Cells were labeled with fixable viability dye eFluor 780 (Thermo Fisher Scientific). Antibodies from Thermo Fisher Scientific included CD19 e780 (clone HIB19), CD14 e780 (clone 61D3), CD16 e780 (clone eBioCB16), OX40 FITC (clone Ber-ACT35), CD69 PeCy7 (clone FN50). Antibodies from Biolegend included: CD45RA BV570 (clone HI100), CXCR5 BV421 (clone J252D4), PD-L1 PE (clone 29E.2A3), CCR7 APC (clone G043H7), CD8a BV650 (clone RPA-T8), CD4 PerCpCy5.5 (clone OKT4). Cells were acquired on a BD Celesta and analyzed using FlowJo Software, version 9.9.6. The frequency of antigen-specific cells was reported by background subtracting the frequency of antigen-specific responses from those of unstimulated cells per donor. Of note, one post-cART specimen did not have sufficient CD4 cells to assess HIV-specific CD4^+^ T cell responses.

### Statistical Analyses

Statistical analyses were performed using GraphPad Prism 9. Non-parametric tests were used in all cases. Wilcoxon and Mann-Whitney U tests were used to compare the magnitude of responses. Fisher exact tests were used to compare the frequency of responders.

## Results

### Study Population

Men who have sex with men (MSM) with HIV were enrolled, from whom blood was taken pre-cART (n=17) and post-cART (n=18) (Table 1). The pre-cART median CD4^+^ T cell count was 606 cells/ul [interquartile range (IQR), 415-718]. The HIV RNA load was 120,000 copies/ml [IQR, 55,087 – 244,480] with a median estimated date of infection of 98 days [IQR, 74-2004] at the time of sample collection. The post-cART median CD4^+^ T cell count was 825 cells/ul [IQR, 563-1025]. The HIV RNA load was below the limit of detection with a median of 667 days [IQR, 495-1388] from beginning cART and a median of 236 days [162-644] from starting cART till an undetectable HIV viral load.

### Detection of HIV-specific CD4^+^ T cells

Using a Class II HIV MP consisting of immunodominant epitopes from the entire HIV proteome, we first tested its ability to detect HIV-specific CD4^+^ T cells in HIV positive and HIV negative samples. In HIV-specific CD4^+^ T cells, there is lower 41BB expression, attributed to T-cell exhaustion from chronic infection which precludes use of 41BB by AIM assay [15]. Additionally, a combination of CD25/OX40 on PBMCs captures more T regulatory cells which may be elevated in chronic infections [10]. We thus identified HIV-specific CD4^+^ T cells by a combination of OX40 and PD-L1 by AIM assay (Fig 1A) [10]. Higher magnitudes of HIV-specific CD4^+^ T cells responses were detected for pre-cART (p=0.00073) and post-cART samples (p=0.0061) compared to HIV negative samples, with a limit of sensitivity of 0.05% (Fig 1B). There was no difference in the proportion of responders detected pre-cART (11/17) and post-cART (10/18) by the Fisher Exact test with a sensitivity of 62.85% and a specificity of 100%, inclusive of paired pre- and post-cART specimens.

**Fig 1.**
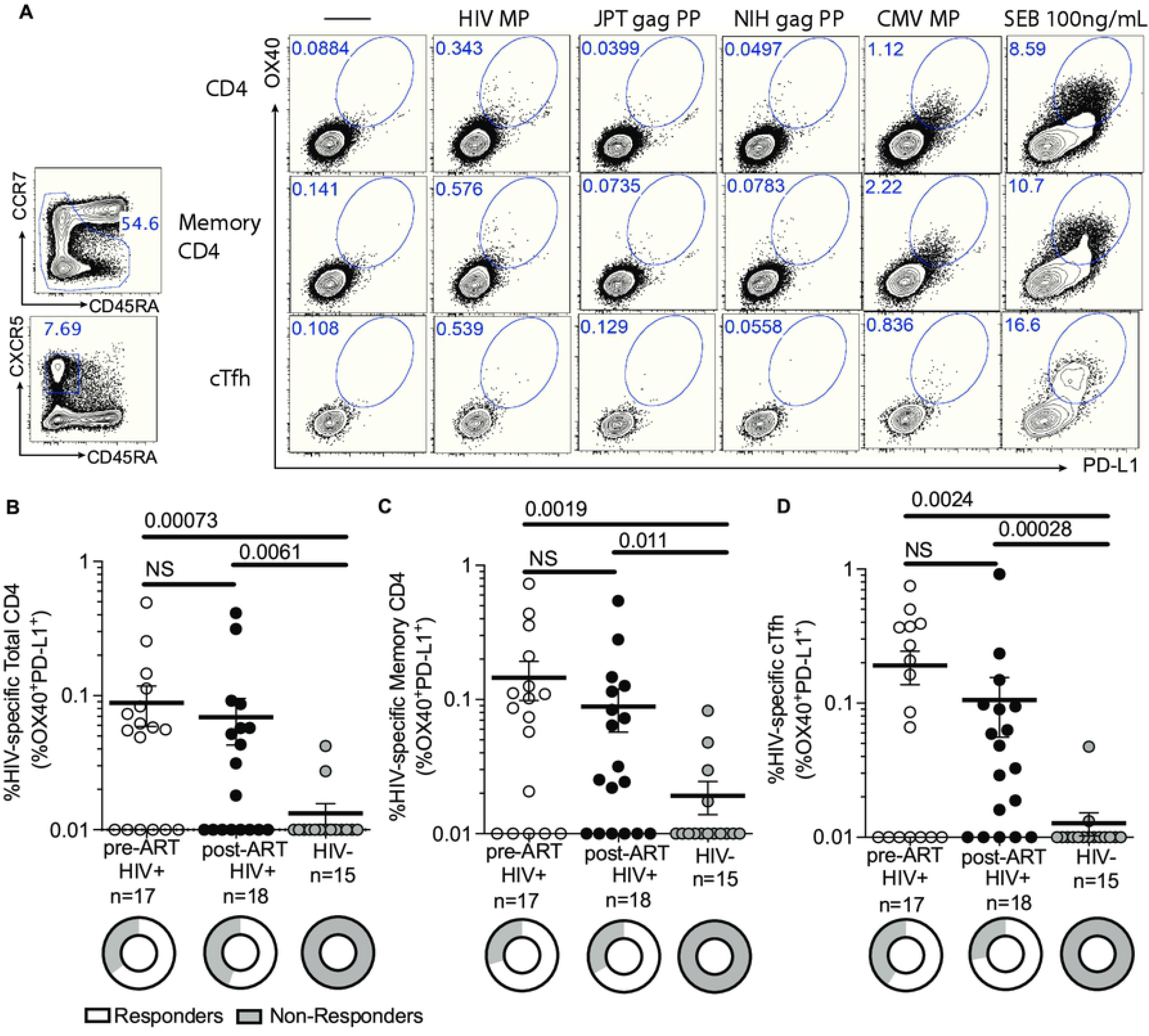
Detection of HIV-specific CD4^+^ T cells. (A) Sample flow cytometry plot showing gating strategy for HIV-specific CD4^+^ T cells based on co-expression of OX40 and PD-L1 on total CD4, memory CD4, and cTfh cells. Specimens from HIV infected donors have a higher frequency of HIV-specific total CD4^+^ T cells, (C) HIV-specific memory CD4^+^ T cells, and (D) HIV-specific cTfh cells compared to HIV uninfected specimens. There was no statistical difference between pre- and post-cART specimens. Data was background subtracted. Statistics by Mann-U Whitney test.

As the memory T cell compartment is important in chronic infection, we quantified memory CD4^+^ T cells. We identified HIV-specific total memory CD4^+^ T cells and circulating T follicular helper (Tfh) cells. We defined total memory CD4^+^ T cells as a combination of central memory + effector memory + T_EMRA_ (Fig 1A). Higher magnitudes of HIV-specific memory CD4^+^ T cell responses were detected in both pre- (p=0.0019) and post-cART HIV-infected individuals (p=0.011, Mann-Whitney) compared to HIV negative individuals (Fig 1C). There was no difference in the proportion of responders pre- (12/17) and post-cART (12/18) by Fisher Exact. Circulating Tfh (cTfh) cells are CD4^+^ T cells which provide help to B cells to promote the development high affinity antibodies [24]. We defined cTfh cells by CCR5^+^CD45RA^-^ CD4^+^ T cells (Fig 1A). Similarly, higher magnitudes of HIV-specific cTfh tells were detected in pre- (p=0.0024) and post-cART HIV-infected individuals (p=0.00028) compared to HIV negative individuals (Fig 1D). There was no difference in the proportion of responders pre- (10/17) and post-cART (13/18) by the Fisher Exact test.

#### Detection of HIV-specific CD8^+^ T cells

Using a Class I HIV MP consisting of immunodominant epitopes from the entire HIV proteome, we tested its ability to detect HIV-specific CD8^+^ T cells in HIV positive and HIV negative samples. By co-expression of CD69 and PD-L1 by AIM assay (Fig 2A), we detected HIV-specific CD8^+^ T cells. The Class I HIV MP detected total CD8^+^ T cells in 11/17 pre- (p=0.0087) and 6/18 post-ART (p=NS) compared to 0/15 HIV negative individuals, with a limit of sensitivity of 0.05% (Fig 2B). For memory CD8^+^ T cells, HIV Class I MP detected 10/17 pre-ART specimens (p=0.021) and 5/18 post-cART specimens (p=NS) compared to 0/15 HIV negative individuals (Fig 2C). There was no difference in the magnitude of CD8^+^ T cell or memory CD8^+^ T cell responses between pre- and post-cART timepoints. The Class I HIV MP had a 48.6% sensitivity and 100% specificity.

**Fig 2.**
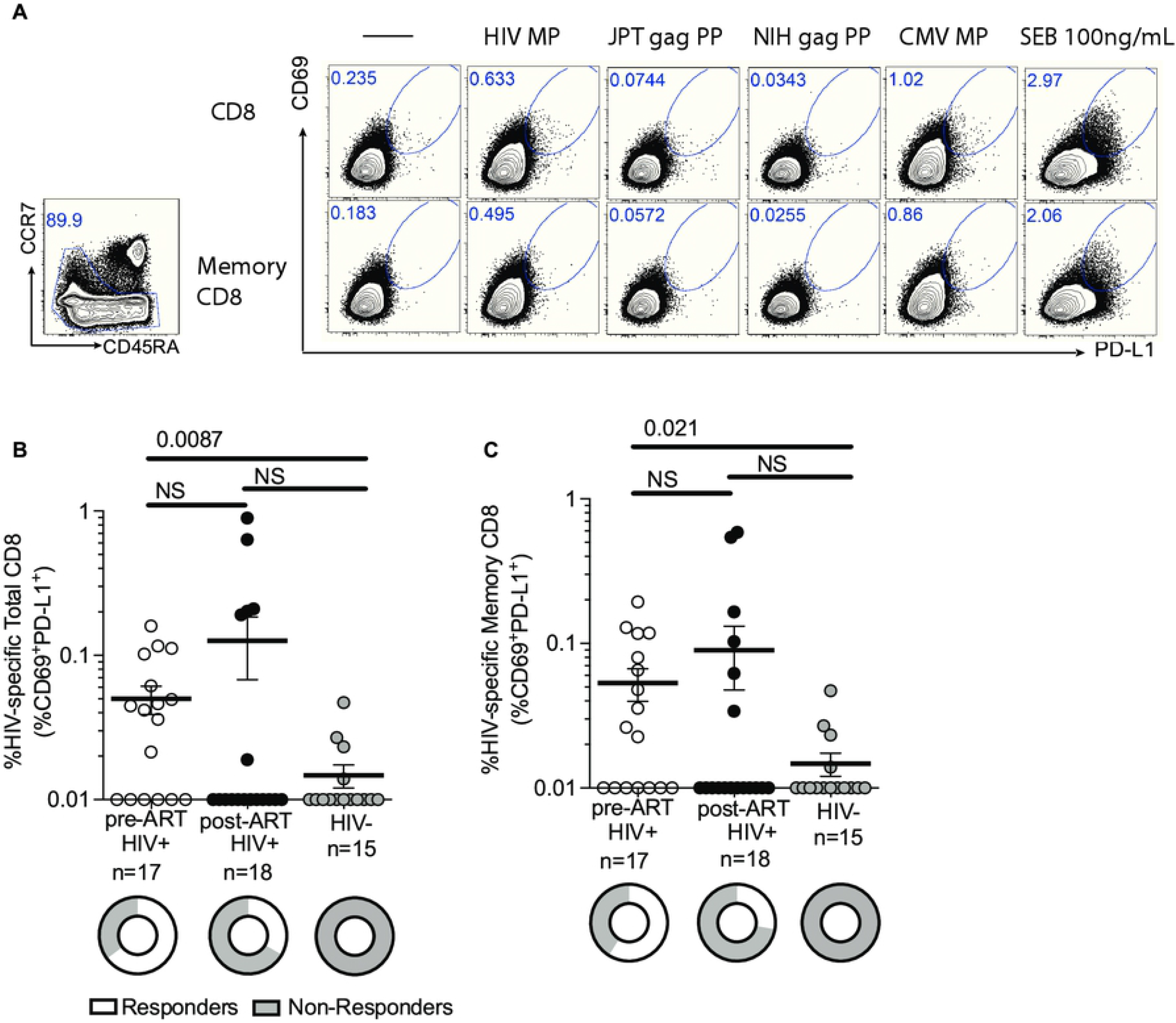
Detection of HIV-specific CD8^+^ T cells. (A) Sample flow cytometry plot showing gating strategy for HIV-specific CD8^+^ T cells based on co-expression of CD69 and PD-L1 on total CD8 and memory CD8 T cells. Specimens from HIV infected donors have a higher frequency of (B) HIV-specific total CD8^+^ T cells and (C) HIV-specific memory CD8^+^ T cells compared to HIV negative specimens. There was no statistical difference between pre- and post-cART specimens. Data was background subtracted. Statistics by Mann-U Whitney test.

#### Comparison of HIV MP to commercially available gag peptide pools

The detection of gag-specific T cells is the most common way to quantify HIV-specific T cells. We therefore compared the Class I and II HIV MP to 2 commercially available gag peptide pools from JPT and NIH. The frequencies of HIV-specific CD4^+^ T cells, memory CD4^+^ T cells, and cTfh were comparable when using the Class II MP, JPT gag, or NIH gag peptide pools (S1A-S1F Figs). The frequencies of HIV-specific total CD8^+^ T cells and memory CD8^+^ T cells were also comparable (S2A-S2D Figs).

**Supplemental Fig 1.**
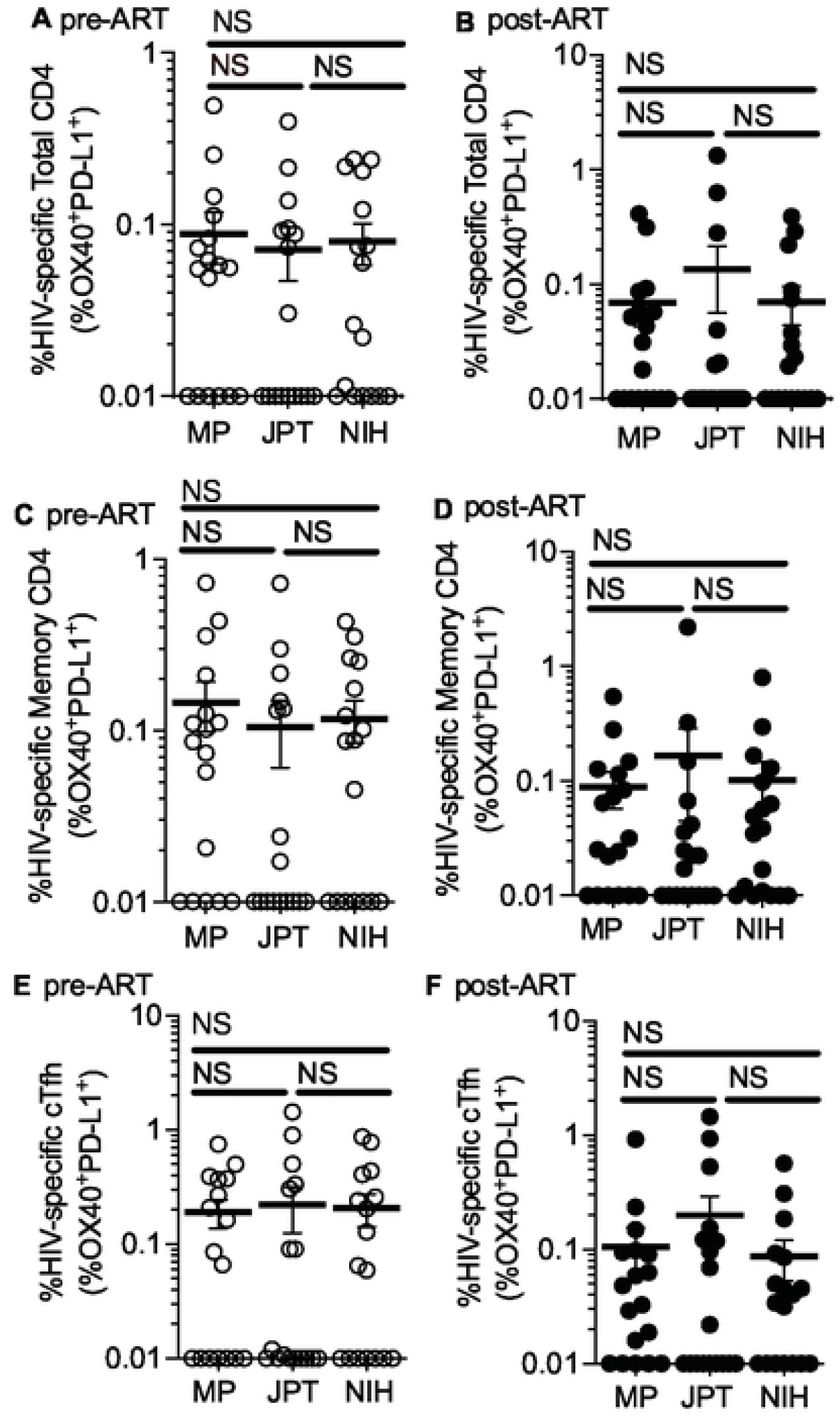
Comparison of HIV-specific CD4^+^ T-cell responses. Comparison of responses between HIV Class II MP, JPT gag, and NIH gag for HIV-specific total CD4^+^ T cells in pre-cART (A) and post-AcRT (B) specimens, HIV-specific memory CD4^+^ T cells in pre-cART and post-cART (D) specimens, and HIV-specific cTfh cells in pre-cART (E) and post-cART (F) specimens. Data was background subtracted. Statistics by Wilcoxon test.

**Supplemental Fig 2.**
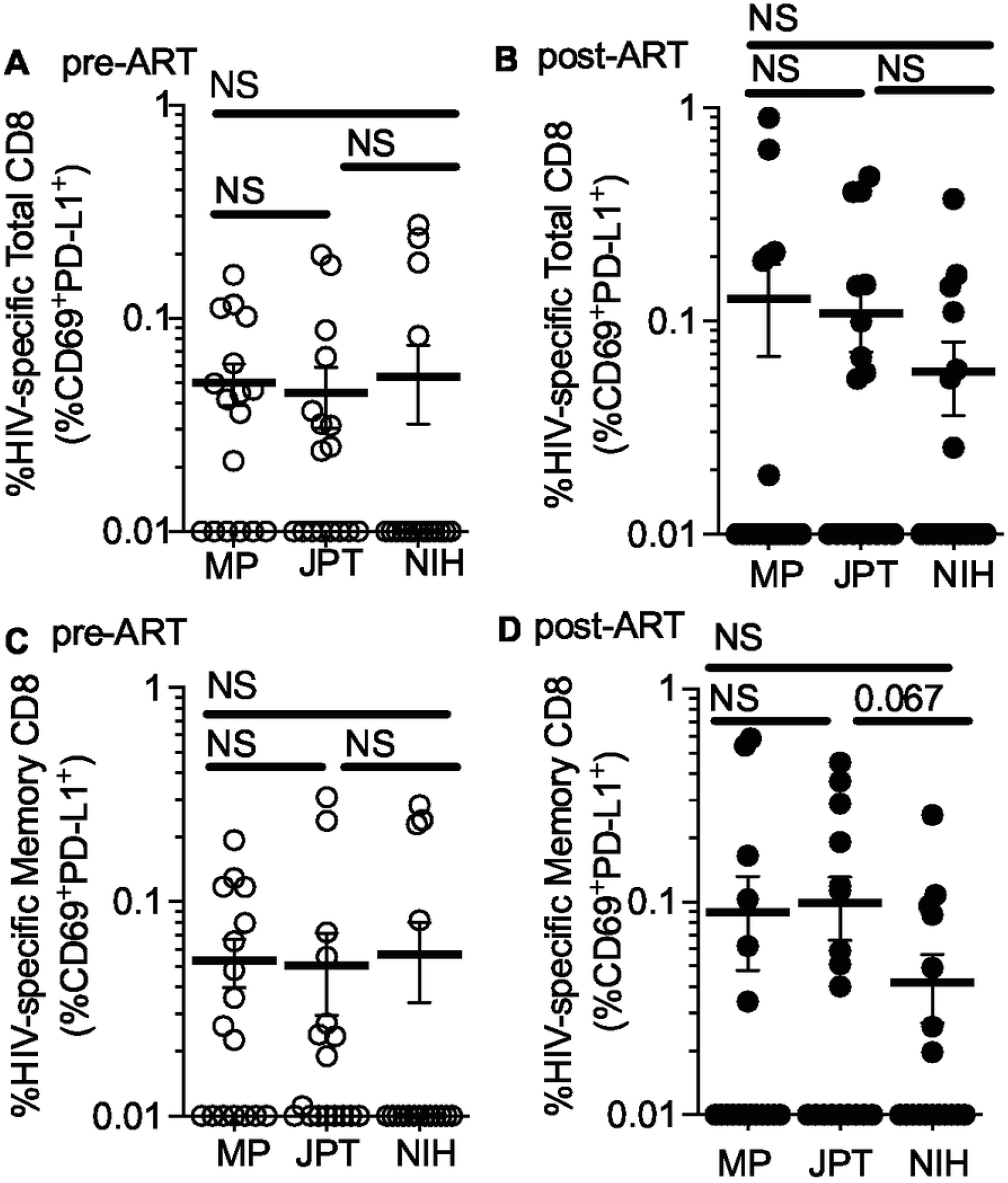
Comparison of HIV-specific CD8^+^ T-cell responses. Comparison of responses between HIV Class I MP, JPT gag, and NIH gag for HIV-specific total CD8^+^ T cells in pre-cART (A) and post-cART (B) specimens and HIV-specific memory CD8^+^ T cells in pre-cART (C) and post-cART (D) specimens. Data was background subtracted. Statistics by Wilcoxon test.

#### Detection of CMV-specific CD4^+^ and CD8^+^ T cells

To demonstrate our ability to detect antigen-specific T cells, as a control, we tested for CMV-specific T cells in these same specimens [25,26]. Using our Class II MP, we were able to detect CMV-specific CD4^+^ T-cells. The magnitudes of CMV-specific CD4^+^ T cells were comparable pre- and post-cART for total CD4^+^ T cells, memory CD4^+^ T cells, and cTfh (S3A-S3C Figs). Using the Class I CMV MP, we additionally tested the magnitudes of CMV-specific CD8^+^ T cells pre- and post-cART. There was no difference between the magnitudes of total and memory CMV-specific CD8^+^ T cell among pre- and post-cART specimens and HIV negative individuals (S3D-S3E Figs).

**Supplemental Fig 3.**
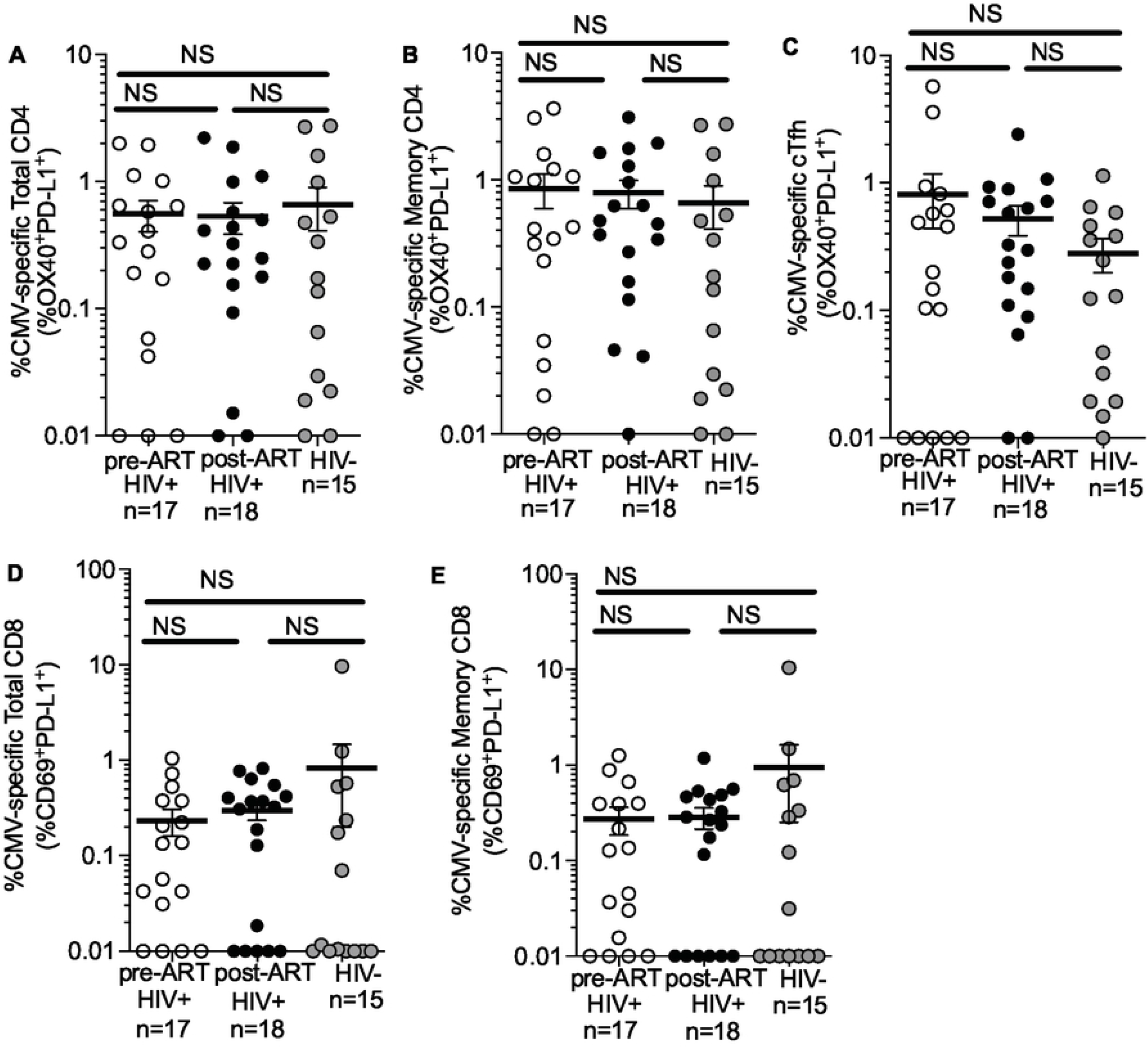
Comparison of CMV-specific responses. Class II MP detects CMV-specific (A) total CD4^+^ T cells, (B) memory CD4^+^ T cells, and (C) cTfh cells. CMV Class I MP detects CMV-specific (D) total CD8^+^ T cells and (E) memory CD8^+^ T cells. Data was background subtracted. Statistics by Mann-U Whitney test.

## Discussion

The HIV MP was designed to encompass all HIV proteins instead of only gag, as other HIV proteins such as nef [17,27,28], pol [29], and tat [30] can elicit HIV-specific T cell responses. Using the Class I and Class II MP reflective of the immunodominant epitopes from the entire HIV proteome, we were able to detect at least either HIV-specific CD4^+^, memory CD4^+^ T, cTfh, CD8^+^ T, or memory CD8^+^ cells in 20/21 (95%) of PWH. Prior studies have demonstrated that 81% of individuals with acute HIV infection had detectable gag-specific CD4^+^ T cells [17] whereas up to 83% had detectable gag-specific CD8^+^ T cells [28]. The one participant in whom we could not detect HIV-specific T-cells had detectable CMV-specific CD8^+^ T cells. For the JPT and NIH gag peptide pools, 19/21 (90%) PWH had either a detectable HIV-specific CD4^+^, memory CD4^+^ T, cTfh, CD8^+^ T, or memory CD8^+^ cells. With regards to detection of HIV-specific T cells in pre-cART (n=17) and post-cART (n=18) specimens, using the HIV MPs, we were unable to detect HIV-specific T cells or their subsets in 1 pre-cART specimen and 4 post-cART specimens. This is consistent with prior studies, in which cART treated individuals have lower frequencies of HIV-specific CD4^+^ and CD8^+^ T cells [4,16,31]. Overall, our HIV Class I and Class II MP were comparable in their ability to detect HIV-specific responses to the gag peptide pools alone, regardless of acute or chronic infection.

PWH are almost universally co-infected with CMV [32]. Our HIV cohort were all CMV IgG seropositive. We were able to detect CMV-specific CD4^+^, memory CD4^+^ T, cTfh, CD8^+^ T, or memory CD8^+^ cells in 21/21 (100%) of PWH.

Limitations of this study include a small sample size comprising entirely of MSM. These individuals comprised both acute infections which were treated early and chronic infections. The frequency of HIV-specific CD4^+^ T cells vary with duration of viremia and therefore antigen exposure, particularly if the patient is suppressed on cART which removes detectable antigen from the bloodstream versus an elite controller [16]. Our cohort consisted of patients with an estimated date of infection ranging from 21 to 3598 days in our pre-cART specimens and time to undetectable viral load ranging from 81 to 1620 days in our post-cART specimens. Moreover, as these participants were enrolled over 2 decades, there were varying cART regimens which differ from today’s first line treatment options. However, we did not observe a clear correlation with T cell responses and cART used.

In summary, we describe an innovative Class I and Class II HIV MP, reflective of the entire HIV proteome, which can be used in future studies to better understand the full repertoire of HIV-specific CD4^+^ and CD8^+^ T cell dynamics. The AIM assay allows for easy quantification of antigen-specific T cells. This methodology permits sorting of live antigen-specific T cells for further functional analyses, allowing for sequencing of CD4^+^ and CD8^+^ subsets, including Tfh cells [8]. These HIV-specific CD4^+^ and CD8^+^ T cells could be sorted and sequenced in hyperacute infections [3] and/or HIV exposed but seronegative individuals [33] and compared to cART treated individuals or elite controllers to assess transcriptomic differences [2].

## Data Availability

Data is available upon request for FACS files. All relevant data are within the manuscript.

---

**Supplemental Table 1.**
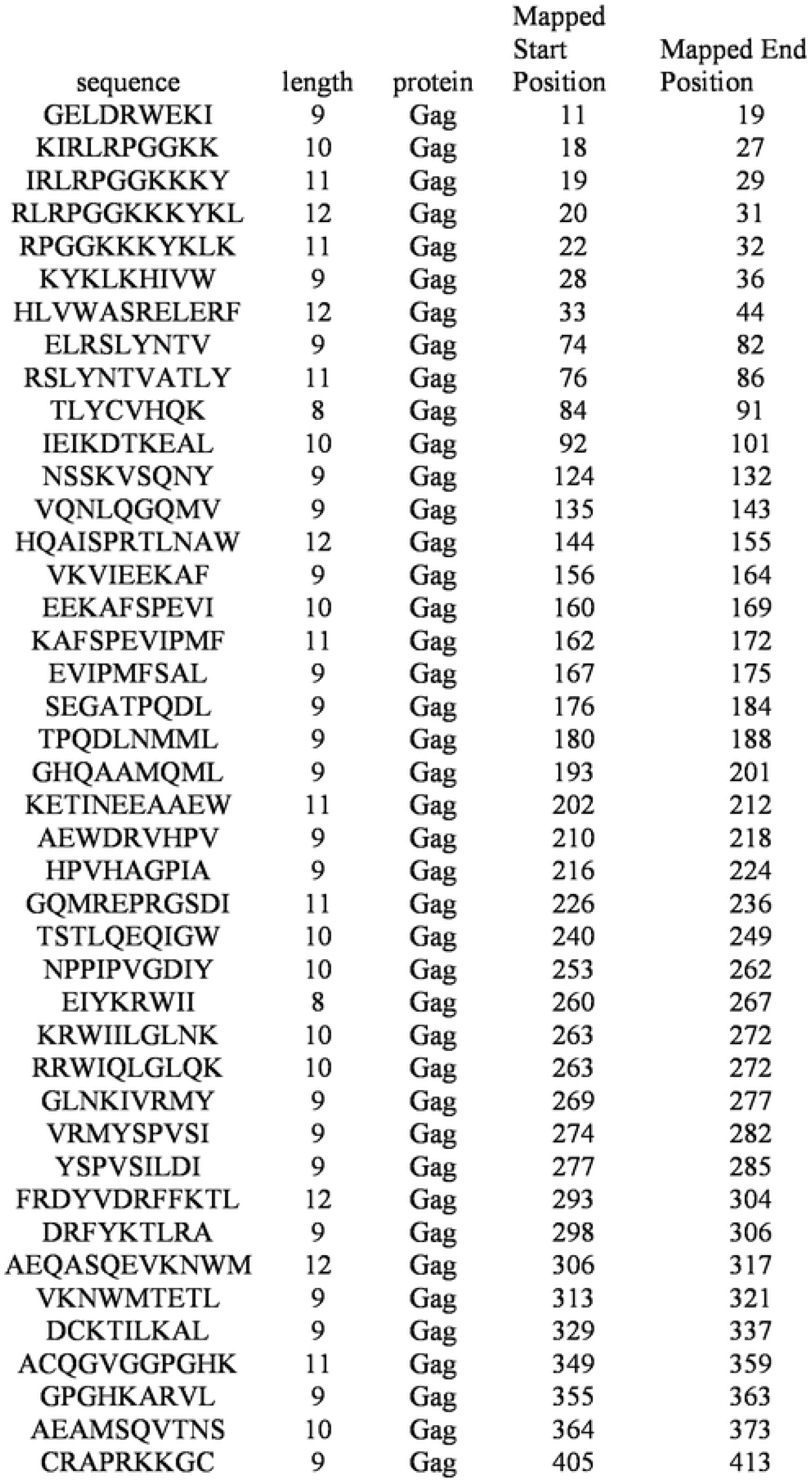
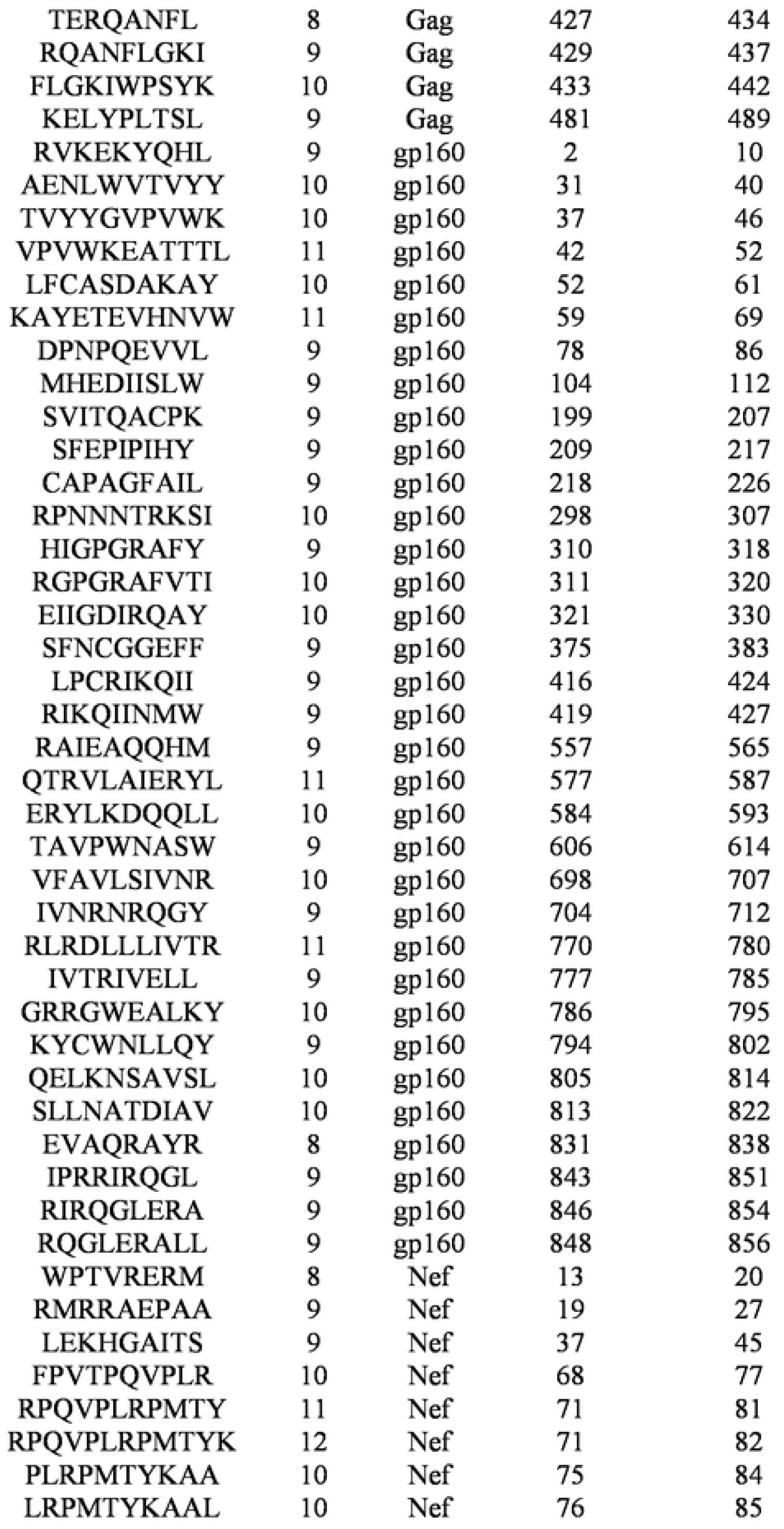
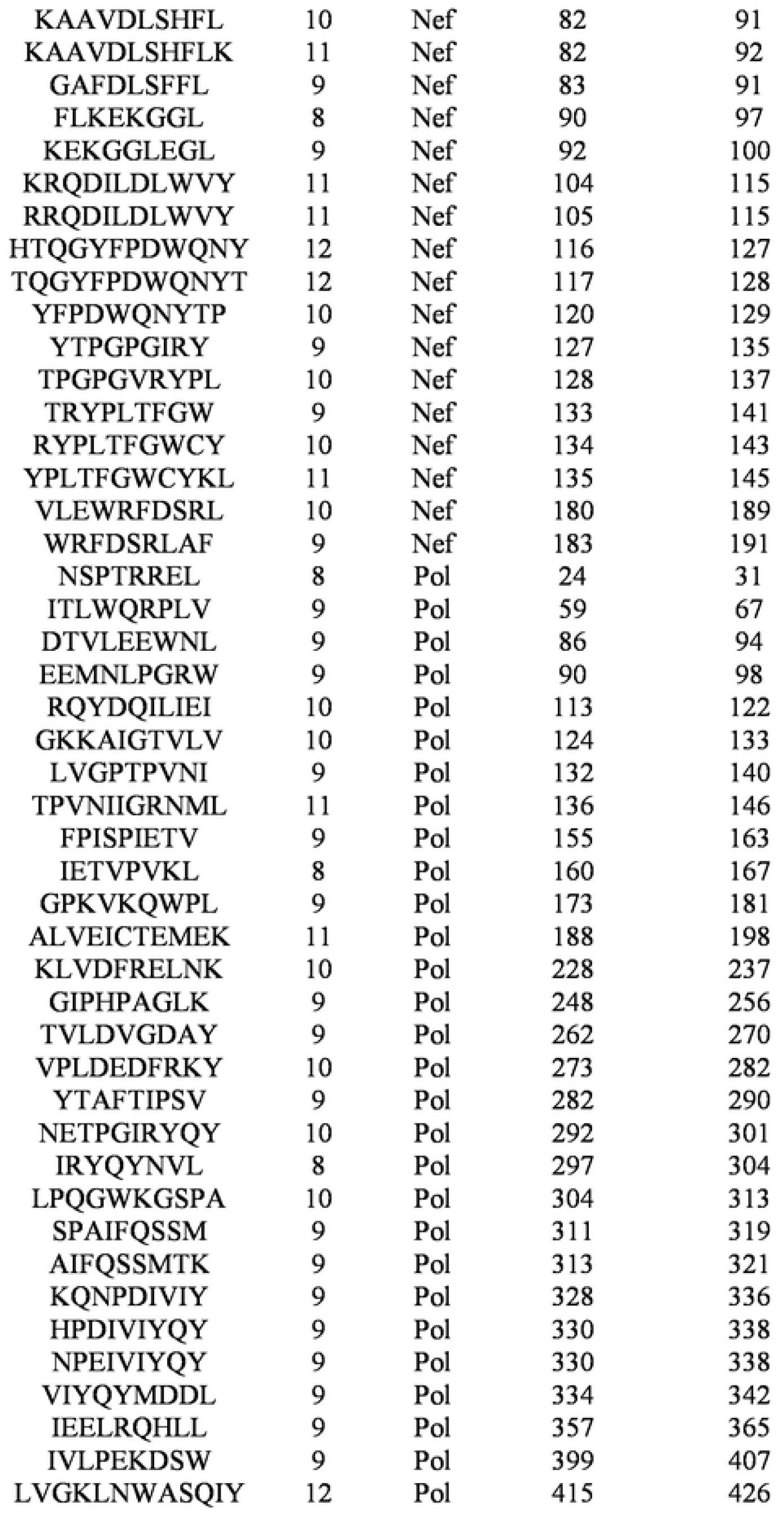
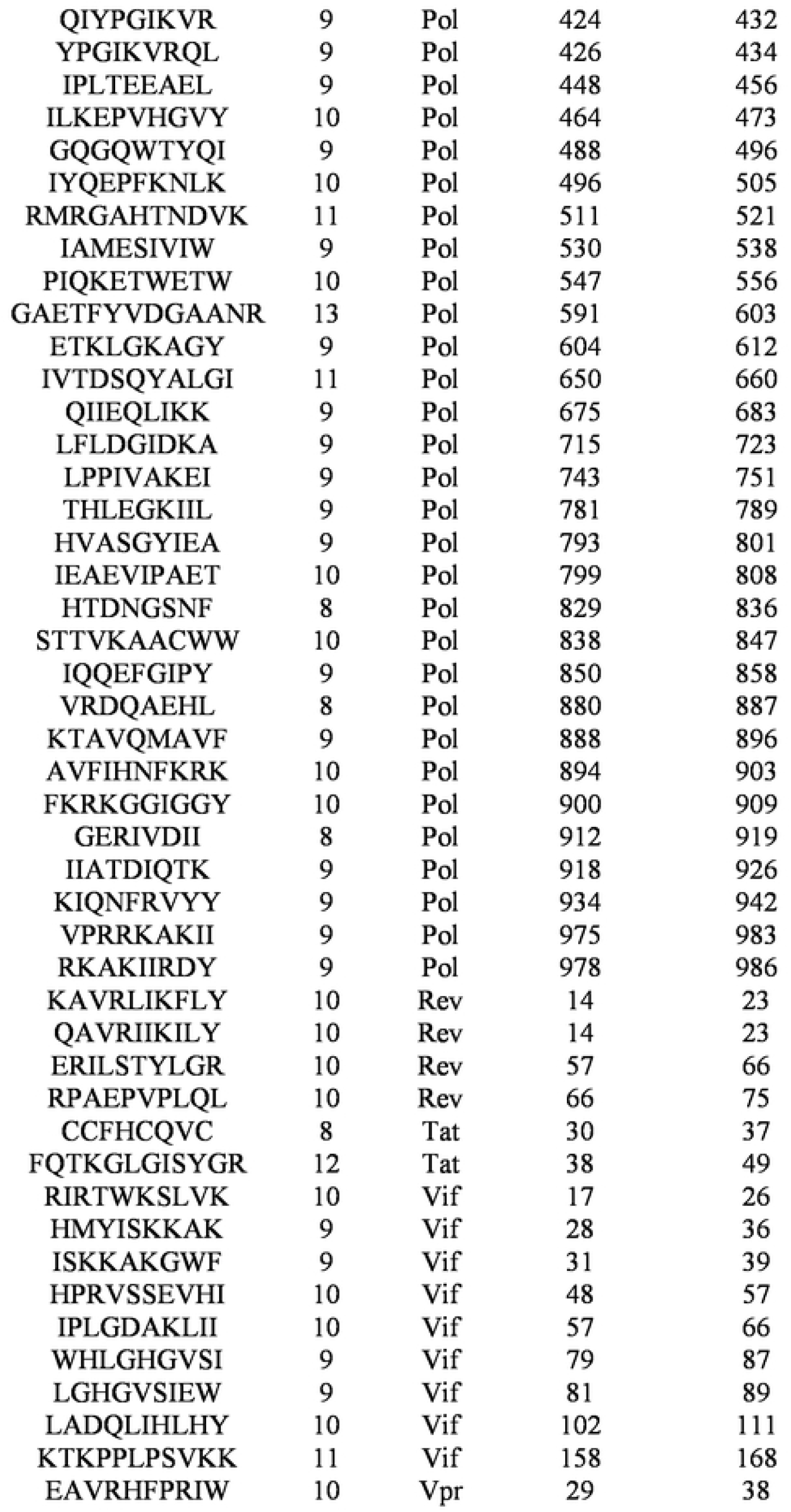
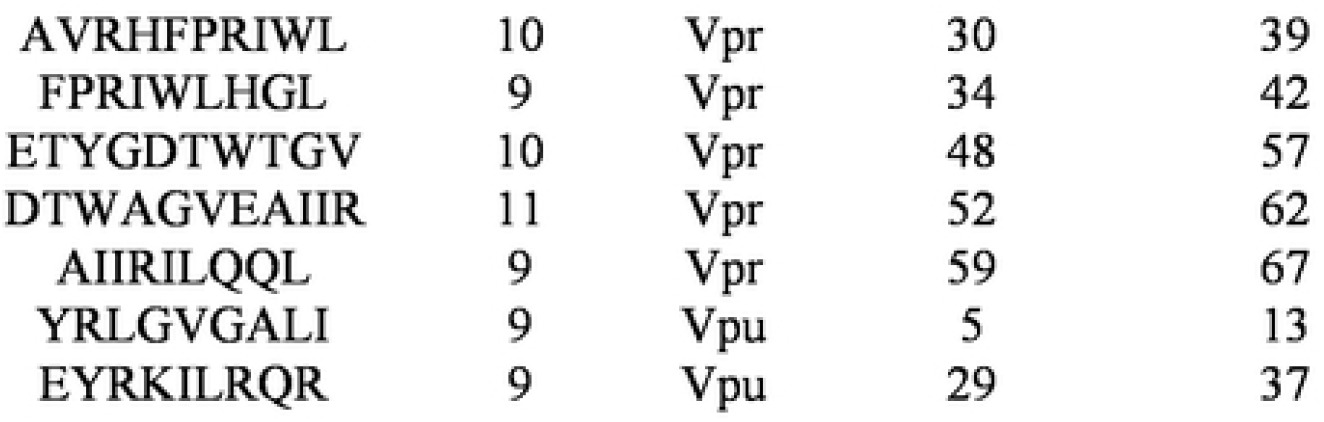
List of HIV class I peptides.

**Supplemental Table 2.**
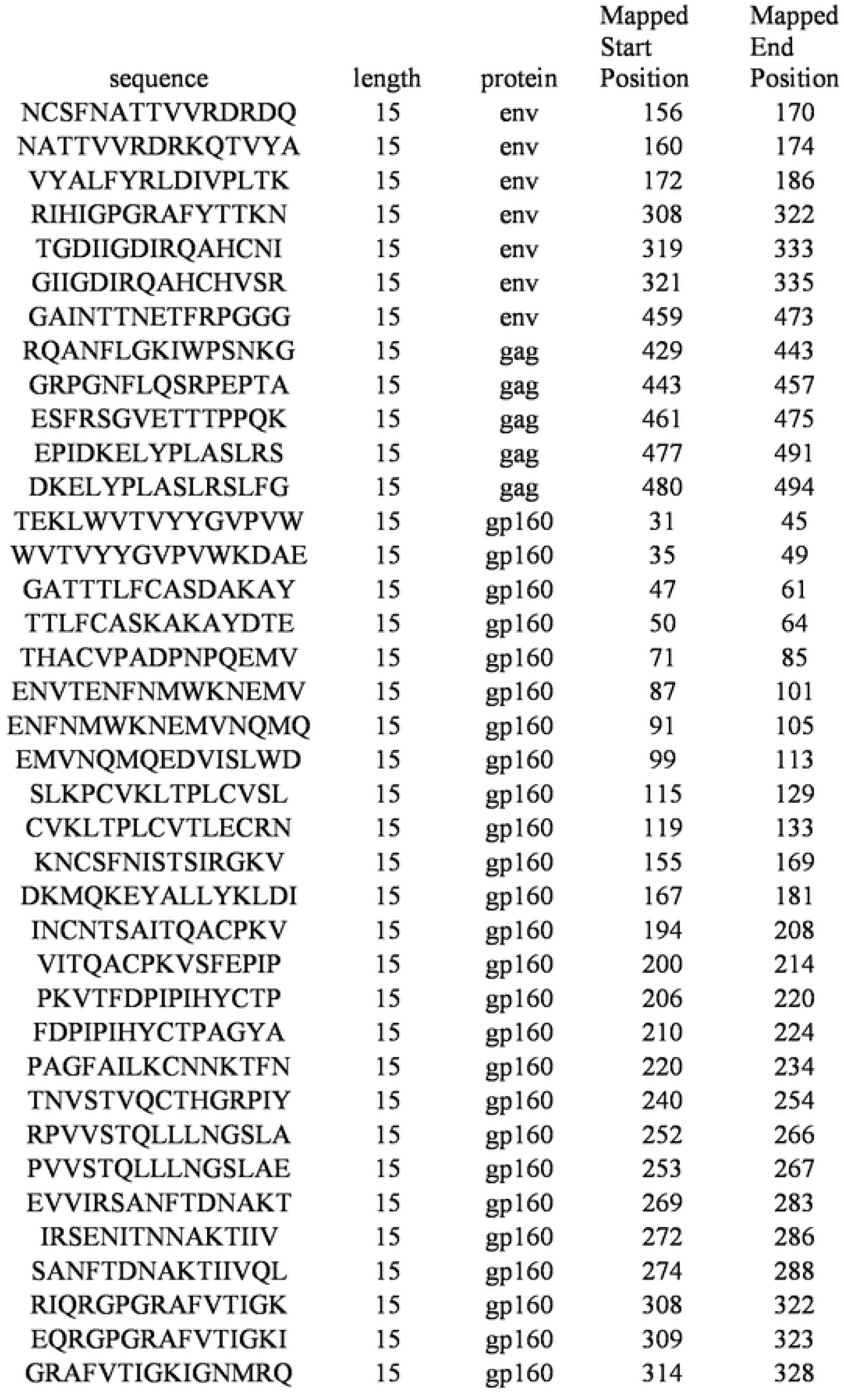
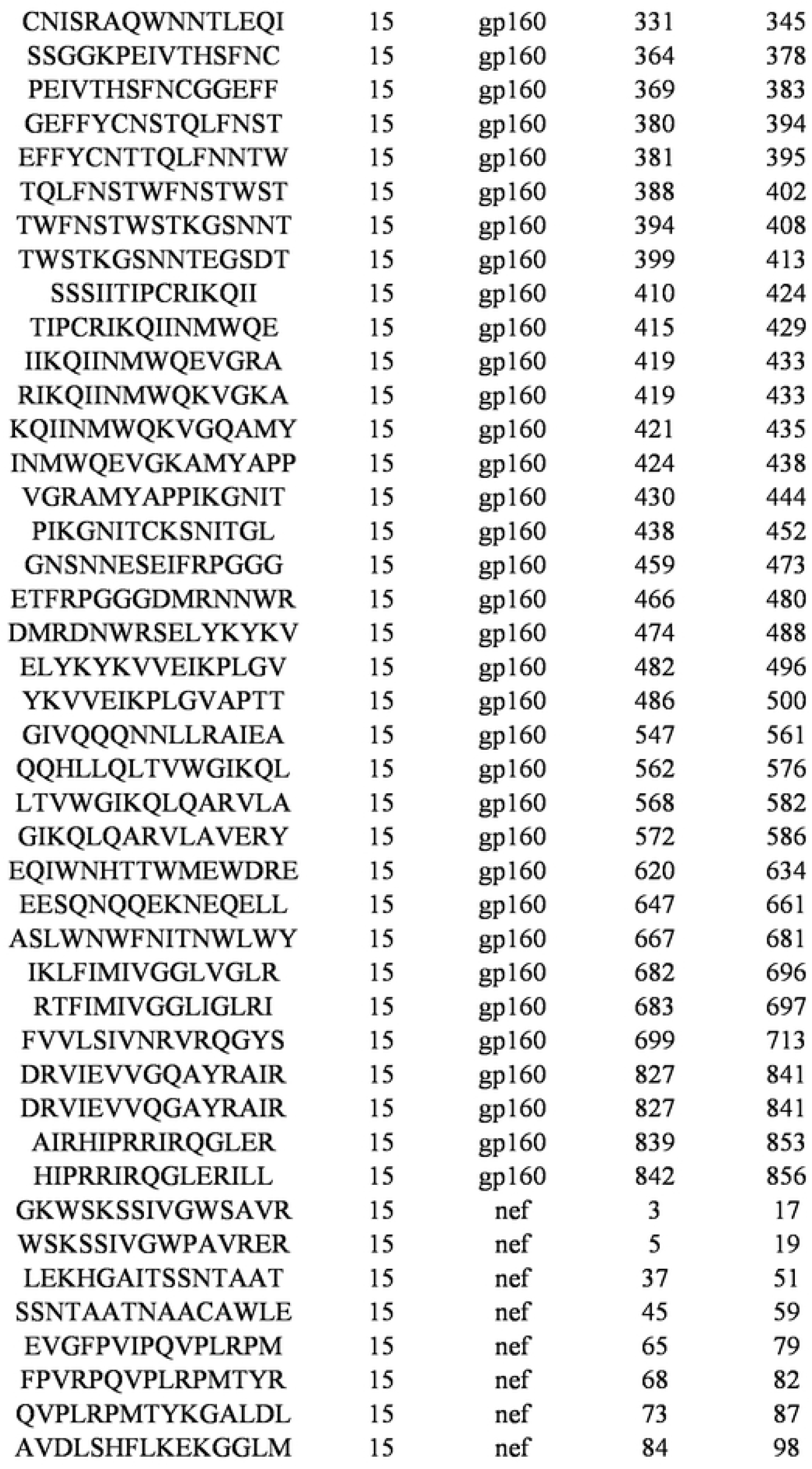
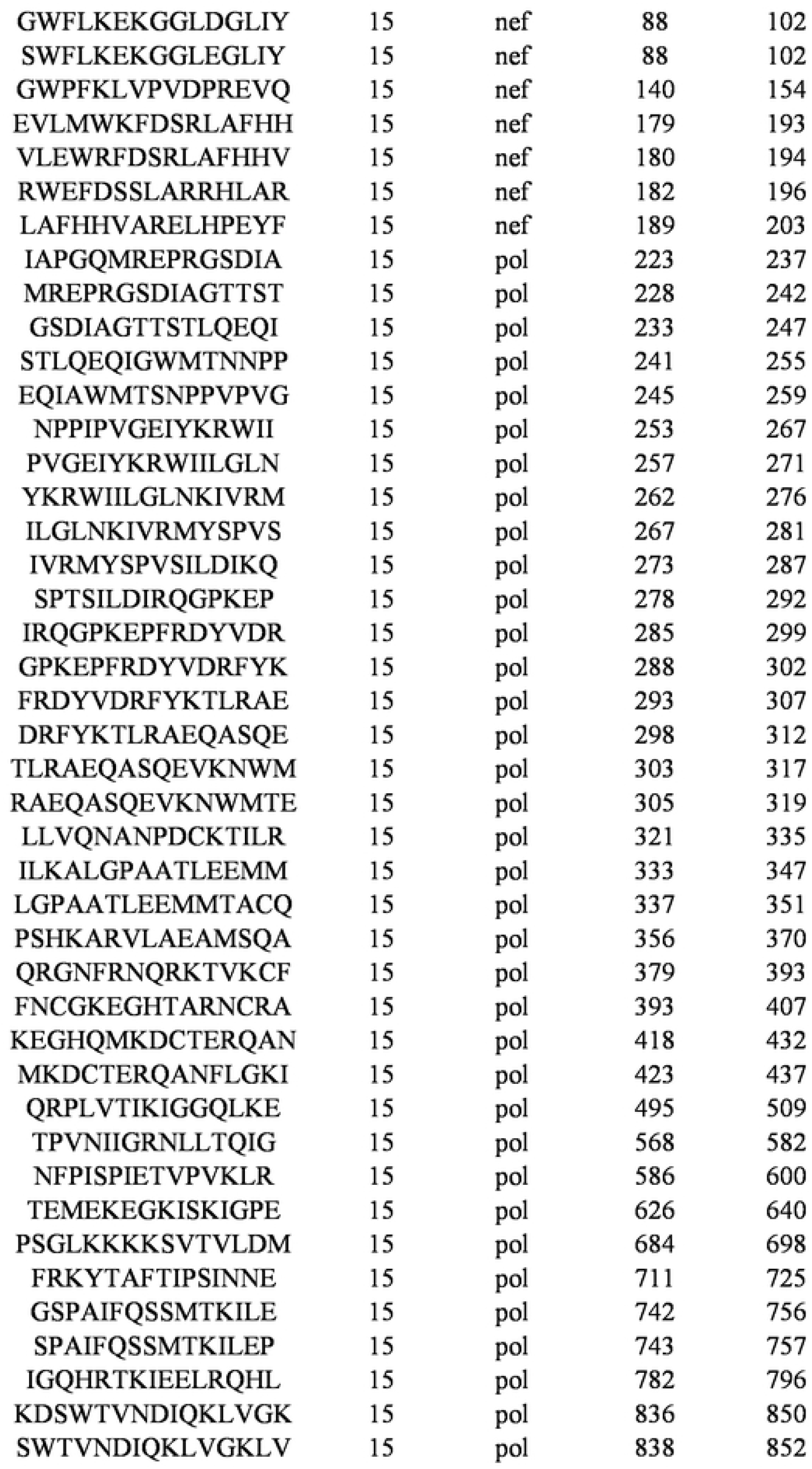
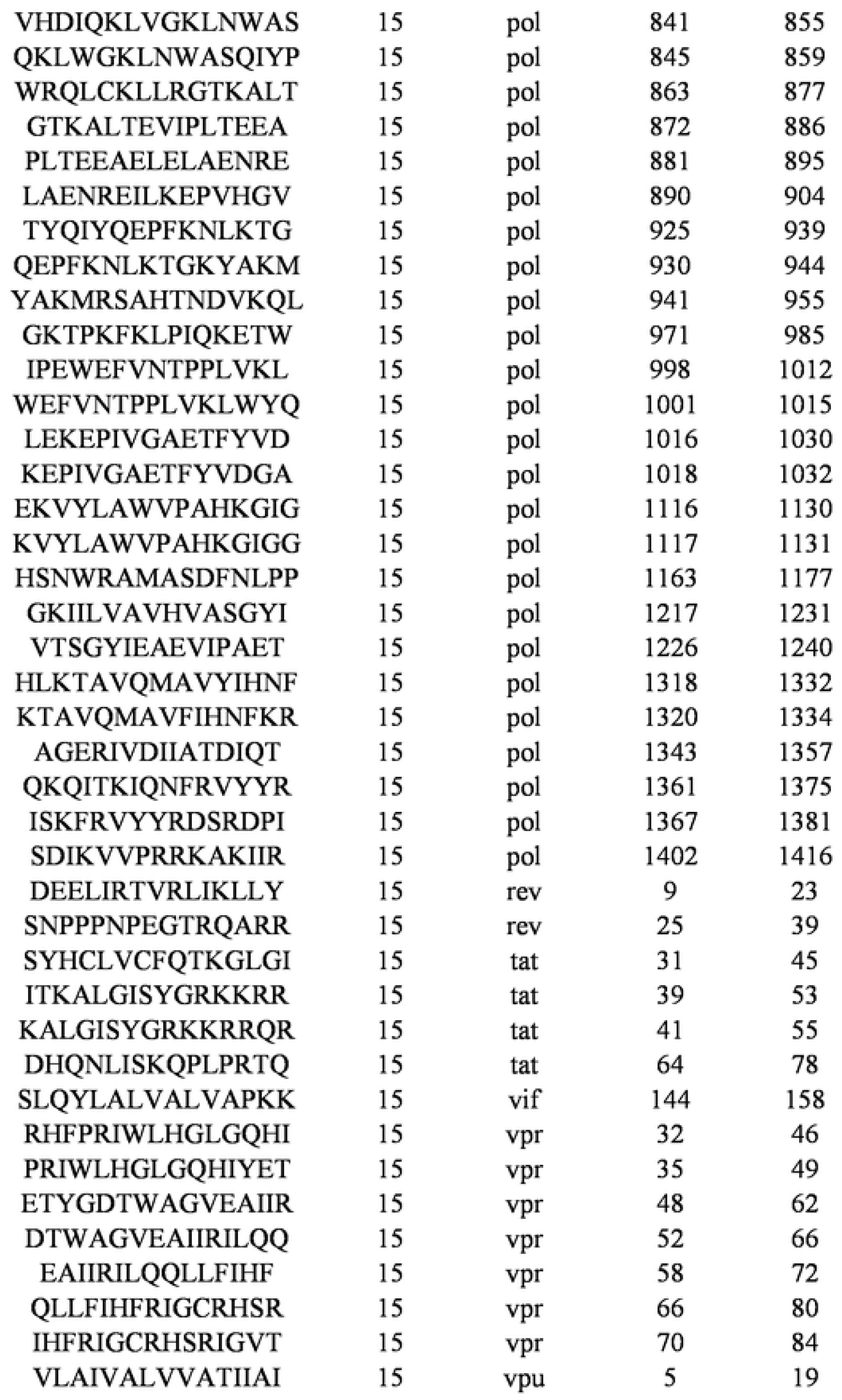
List of HIV class II peptides.

